# The Healthy Brain Initiative (HBI): A prospective cohort study protocol

**DOI:** 10.1101/2023.09.21.23295908

**Authors:** Lilah M. Besser, Stephanie Chrisphonte, Michael J. Kleiman, Deirdre O’Shea, Amie Rosenfeld, Magdalena Tolea, James E. Galvin

## Abstract

**Background:** The Health Brain Initiative (HBI), established by University of Miami’s Comprehensive Center for Brain Health (CCBH), follows racially/ethnically diverse older adults without dementia living in South Florida. With dementia prevention and brain health promotion as an overarching goal, HBI will advance scientific knowledge by developing novel assessments and non-invasive biomarkers of Alzheimer’s disease and related dementias (ADRD), examining additive effects of sociodemographic, lifestyle, neurological and biobehavioral measures, and employing innovative, methodologically advanced modeling methods to characterize ADRD risk and resilience factors and transition of brain aging.

**Methods:** HBI is a longitudinal, observational cohort study that will follow 500 deeply-phenotyped participants annually to collect, analyze, and store clinical, cognitive, behavioral, functional, genetic, and neuroimaging data and biospecimens. Participants are ≥50 years old; have no, subjective, or mild cognitive impairment; have a study partner; and are eligible to undergo magnetic resonance imaging (MRI). Recruitment is community-based including advertisements, word-of-mouth, community events, and physician referrals. At baseline, following informed consent, participants complete detailed web-based surveys (e.g., demographics, health history, risk and resilience factors), followed by two half-day visits which include neurological exams, cognitive and functional assessments, an overnight sleep study, and biospecimen collection. Structural and functional MRI is completed by all participants and a subset also consent to amyloid PET imaging. Annual follow-up visits repeat the same data and biospecimen collection as baseline, except that MRIs are conducted every other year after baseline.

**Ethics and expected impact:** HBI has been approved by the University of Miami Miller School of Medicine Institutional Review Board. Participants provide informed consent at baseline and are re-consented as needed with protocol changes. Data collected by HBI will lead to breakthroughs in developing new diagnostics and therapeutics, create comprehensive diagnostic evaluations, and provide the evidence base for precision medicine approaches to dementia prevention with individualized treatment plans.

## INTRODUCTION

Alzheimer’s disease and related dementias (ADRD) affect approximately 55 million individuals worldwide, and the prevalence is projected to rise with the expected surge in the population of older adults over the next few decades (1). ADRD are neurodegenerative diseases that result in cognitive and functional decline, with initial symptoms varying based on etiology. For example, Alzheimer’s disease (AD), which involves the accumulation of amyloid plaques and tau tangles throughout the brain, typically begins with episodic memory problems experienced during older age (e.g. ≥65 years) (2). In contrast, Lewy body dementia (LBD), which is associated with the accumulation of alpha-synuclein protein or Lewy bodies in the brain, usually presents as having motor symptoms, cognitive fluctuations, rapid eye movement (REM) sleep behavior disorder, and/or visual hallucinations starting at age 50 or older (3). Some recent advances have been made in clinical trials, such as monoclonal antibody treatments (e.g., aducanumab, lecanumab, donanemab), which have been shown to remove amyloid plaques and slow cognitive decline among individuals with early AD (4). These monoclonal antibody treatments have been associated with amyloid-related imaging abnormalities and therefore require careful monitoring with serial Magnetic Resonance Imaging (MRI) (5) that is costly, time-consuming, and burdensome. However, no treatments are presently available to prevent incipient ADRD. Thus, while much research effort remains directed at drug development focused on removing ADRD pathology to slow or halt cognitive decline amongst symptomatic individuals, there is still a great need for studies to identify at-risk individuals and develop primary and secondary prevention strategies.

Older age, familial dominant genetic mutations (e.g., APP, PSEN1, PSEN2 for AD), and genetic risk factors (e.g., ≥1 apolipoprotein e4 allele) contribute significantly to an individual’s vulnerability toward developing ADRD (6). Beyond these non-modifiable risk factors, 12 modifiable risk factors were identified by the Lancet Commission in 2020 as significant contributors to dementia risk based on a review of the extant literature (7). These risk factors, which were estimated to account for approximately 40% of dementias worldwide, include less education, smoking, obesity, hearing impairment, depression, physical inactivity, hypertension, diabetes, low social contact, traumatic brain injury (TBI), alcohol consumption, and air pollution exposure. The estimates of population attributable fraction (PAF) from the Lancet commission have been modified over time (from 2017 (8) to 2020 (7)) to add TBI, air pollution, and alcohol use), and will likely be modified again as evidence accumulates for additional risk factors that have not been adequately studied to date. In addition, the PAF estimates were based on a sample in the United Kingdom, which might have different risk factors that regions with differing diversity by factors such as racial/ethnic group. Altogether, the 12 identified risk factors represent an initial list of important dementia risk factors that will likely be expanded based on evidence from new studies and more diverse cohorts.

Recently, clinical and epidemiological studies have shifted to not only focus on risk factors for developing dementia, but also on the unique factors that impart resistance against the development of ADRD neuropathology or resilience against cognitive decline in the face of underlying neuropathology (9). This shift in orientation parallels an increased research interest in brain health, which is the “preservation of optimal brain integrity and mental and cognitive function at a given age in the absence of overt brain diseases that affect normal brain function” (10). Pathology associated with ADRD can begin decades before initial symptom presentation, during the midlife period (11). Use of biomarkers such as those ascertained from cerebrospinal fluid and blood (e.g., amyloid beta, p-Tau) is particularly important to help identify protective factors associated with lowering ADRD risk when symptoms have not yet manifested but pathology may be present. While growing, research is limited on risk and protective factors in relation to ADRD biomarkers with an eye toward cognitive reserve and resilience. In addition, research needs to expand to investigate neurobehavioral biomarkers strongly predictive of ADRD but that are less invasive than the established biomarkers that require magnetic resonance imaging (MRI), positron emission tomography (PET), cerebrospinal fluid (CSF), or blood or skin biopsy collection (12), and that could be applied in clinical practice.

The NIA-AA research ATN framework emphasizes the incorporation of biomarker measures of amyloid, tau, and neurodegeneration/neuronal injury, providing an opportunity to classify individuals across the AD spectrum (13). For instance, abnormal amyloid levels have been consistently shown to predict subsequent cognitive decline as well as progression to mild cognitive impairment and dementia (14–17). Risk of future decline among amyloid+ normal controls is particularly high in those that also have abnormal tau (14, 18) or neurodegeneration (19–21). Combining information across ATN dimensions allows identification of clinically normal individuals who are in the earliest stages of the AD spectrum (“preclinical AD”). In addition, ATN dimensions can be applied in differential diagnosis (13, 21–23), assessing target engagement during clinical trials (24), and determine presence of other pathologies such as Lewy bodies (13, 25). Overall, the ATN framework provides a new perspective for studies of risk for and resilience against ADRD. Recently, revisions have been proposed for the ATN framework to incorporate other markers of neuronal injury, neurodegeneration, and coexisting pathologies (e.g., vascular lesions, Lewy bodies).

The gaps in the literature on ADRD risk and protective factors, cognitive reserve, and resilience are particularly apparent for minoritized and disadvantaged populations (e.g., racial/ethnic minoritized and lower socioeconomic status (SES) groups) (26). It is critical for future work on brain health promotion and ADRD prevention to focus on protective factors for these populations, who also appear to be at higher risk of developing ADRD (6). For instance, Black and Hispanic individuals are 1.5 to 2 times more likely to develop ADRD than White individuals, and risk is highest amongst those with a combination of low education and low lifetime occupational attainment (27). Factors that can be targeted via intervention to reduce ADRD risk may differ between socially disadvantaged and advantaged individuals, and new cohorts are needed with diverse populations including understudied immigrant populations.

Studies on unique factors promoting cognitive reserve and resilience (beyond known risk factors) that can be targeted via intervention to prevent ADRD remain limited. The Health Brain Initiative (HBI) cohort was established by University of Miami’s Comprehensive Center for Brain Health (CCBH) to fill this gap and advance beyond the studies of healthy brain aging and ADRD research to date. HBI will follow a racially/ethnically diverse cohort of ≥50-year-olds with normal cognition, subjective cognitive complaints, mild cognitive impairment or very mild dementia from the South Florida region. With ADRD prevention and brain health promotion as an overarching goal, HBI will advance scientific knowledge on established and novel ADRD biomarkers, examine additive effects of multiple factors (e.g., sociodemographics and lifestyle) on risk, reserve, and resilience, and employ innovative and methodologically advanced methods to characterize risk/resilience factors and disease progression.

## MATERIALS AND METHODS

### Study aims, design, and setting

The HBI is a longitudinal, observational cohort study that will prospectively collect, analyze, maintain, and store clinical, cognitive, behavioral, functional and neuroimaging data and biospecimens from research participants, to allow a characterization of transitions in cognitive status, studying risk and protective factors for cognitive and functional decline, and creation of a repository of data for future research questions. The cohort will have representative sampling by sex, race and ethnicity for normal cognition (NIA-AA Stage 1), subjective memory complaints (Stage 2), MCI (Stage 3), or mild ADRD (Stage 4) according to the ATN framework (13). HBI’s specific aims are to:

1. Characterize novel approaches for detection of healthy brain aging and ADRD and examine their relationship to ADRD biomarkers and ADRD-related outcomes (Aim 1);
2. Examine potential sociodemographic, biological, genetic, health and lifestyle additive effects on the presence and accumulations of ADRD pathology detected via biomarkers that may confer risk and resilience to cognitive decline (Aim 2);
3. Determine whether potentially modifiable risk factors across the life course buffer the impact of brain pathology on cognitive decline and thus can serve as intervention targets (Aim 3);
4. Use machine learning and other advanced analytic techniques to explore trajectories of disease progression by baseline health, lifestyle, and other risk factors (Aim 4); and,
5. Conduct study aims with a racially/ethnically and socioeconomically diverse cohort that will allow for examination of disparities in biomarkers, ADRD outcomes, and risk/protective factors (Aim 5).

HBI will recruit a longitudinal cohort of 500 participants living in the racially/ethnically diverse South Florida region, primarily Palm Beach and Broward counties. Multiple grant-funded studies will use the standardized HBI protocol for data collection and thus contribute to the target recruitment goals for HBI (e.g., NIH R01AG071514, R01AG071514S1, R01NS101483, R01NS101483S1, RF1AG075901, and R56AG074889).

Recruitment will occur through a variety of sources, including advertisements, word-of-mouth, community lectures, health fairs, community events, educational conferences, physician referrals, and working with community partners. HBI has an IRB-approved cross-sectional survey of risk and resilience factors that can be used to identify eligible individuals. Individuals who call the center will be provided with IRB-approved literature regarding the study objectives and inclusion/exclusion criteria. A full-time bilingual recruitment coordinator will work in conjunction with the research staff to organize and conduct recruitment activities.

Annual visits will be conducted at University of Miami’s Comprehensive Center for Brain Health (CCBH) in Boca Raton, Florida. The visits will involve both the participant and their study partner, who is typically a spouse or adult child who provides independent, collateral information on the participant. Our prior research has demonstrated the importance of the caregiver-patient dyad to longitudinal outcomes (28–31).

### Inclusion and exclusion criteria

The target sample of 500 individuals to be followed longitudinally will include those who:

- Are ≥50 years old,
- Have no cognitive impairment, subjective cognitive impairment (SCI), mild cognitive impairment (MCI), or any cause of dementia or neurodegenerative disease at the mild stage (Clinical Dementia Rating (CDR®)≤1),
- Have a study partner; and,
- Are medically eligible to undergo MRI.

Individuals will be excluded if they:

- Do not consent for data/specimen storage,
- Have already been diagnosed with moderate or severe ADRD (Clinical Dementia

Rating≥2 (32)),

- Cannot contribute clinical, cognitive, behavioral, or functional data; or,
- Have significant medical illness that could affect participation in imaging or obfuscate relevant cognitive outcomes (e.g., metastatic cancer, primary Axis I disorder such as schizophrenia, Axis II personality disorder, unstable diabetes or hypertension, or substance abuse within the past 5 years).

### Procedures

#### Baseline visit

Participants and their study partners will be consented at the baseline visit, and then reconsented at annual follow-ups if needed. Following informed consent, the participants will complete a battery of questionnaires collecting information such as demographics, medical history to identify possible risk factors for memory problems (e.g., high blood pressure, strokes, heart disease, diabetes, other potentially relevant comorbidities), lifestyle activities, and resilience and vulnerability factors (Tables 1 and 2).

**Table 1.**
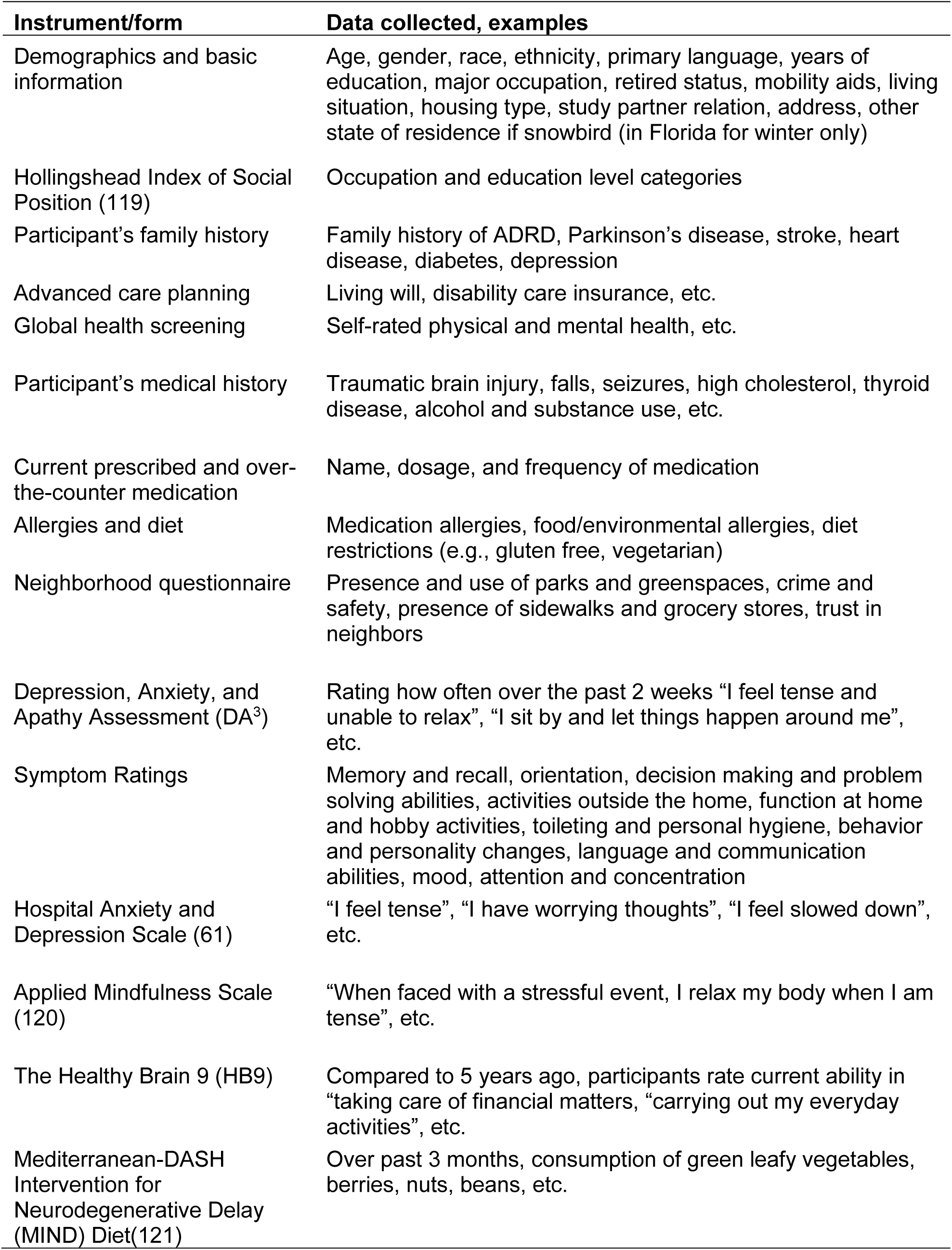
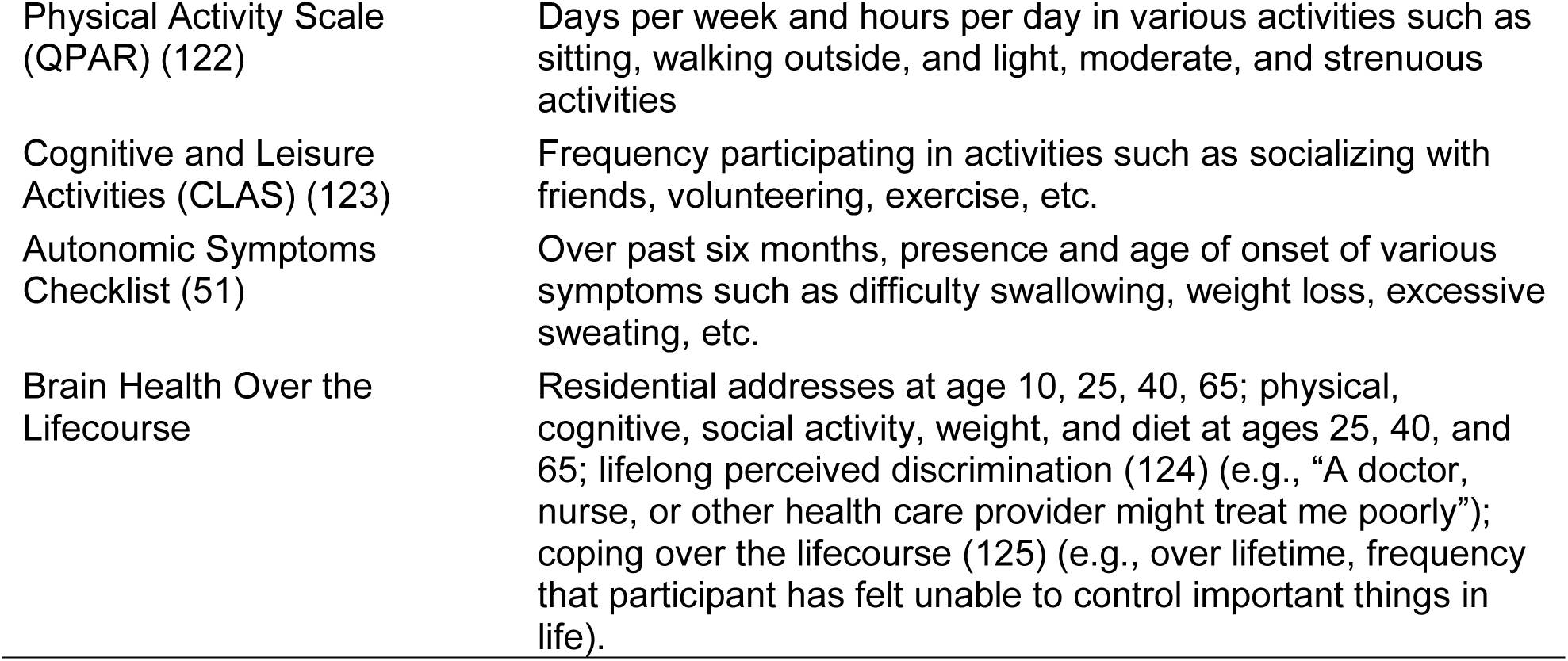
Participant questionnaires.

| **Instrument/form** | **Data collected, examples** |
| --- | --- |
| Demographics and basic information | Age, gender, race, ethnicity, primary language, years of education, major occupation, retired status, mobility aids, living situation, housing type, study partner relation, address, other state of residence if snowbird (in Florida for winter only) |
| Hollingshead Index of Social Position (119) | Occupation and education level categories |
| Participant's family history | Family history of ADRD, Parkinson's disease, stroke, heart disease, diabetes, depression |
| Advanced care planning | Living will, disability care insurance, etc. |
| Global health screening | Self-rated physical and mental health, etc. |
| Participant's medical history | Traumatic brain injury, falls, seizures, high cholesterol, thyroid disease, alcohol and substance use, etc. |
| Current prescribed and over-the-counter medication | Name, dosage, and frequency of medication |
| Allergies and diet | Medication allergies, food/environmental allergies, diet restrictions (e.g., gluten free, vegetarian) |
| Neighborhood questionnaire | Presence and use of parks and greenspaces, crime and safety, presence of sidewalks and grocery stores, trust in neighbors |
| Depression, Anxiety, and Apathy Assessment (DA<sup>3</sup>) | Rating how often over the past 2 weeks "I feel tense and unable to relax", "I sit by and let things happen around me", etc. |
| Symptom Ratings | Memory and recall, orientation, decision making and problem solving abilities, activities outside the home, function at home and hobby activities, toileting and personal hygiene, behavior and personality changes, language and communication abilities, mood, attention and concentration |
| Hospital Anxiety and Depression Scale (61) | "I feel tense", "I have worrying thoughts", "I feel slowed down", etc. |
| Applied Mindfulness Scale (120) | "When faced with a stressful event, I relax my body when I am tense", etc. |
| The Healthy Brain 9 (HB9) | Compared to 5 years ago, participants rate current ability in "taking care of financial matters, "carrying out my everyday activities", etc. |
| Mediterranean-DASH Intervention for Neurodegenerative Delay (MIND) Diet(121) | Over past 3 months, consumption of green leafy vegetables, berries, nuts, beans, etc. |
| Physical Activity Scale (QPAR) (122) | Days per week and hours per day in various activities such as sitting, walking outside, and light, moderate, and strenuous activities |
| Cognitive and Leisure Activities (CLAS) (123) | Frequency participating in activities such as socializing with friends, volunteering, exercise, etc. |
| Autonomic Symptoms Checklist (51) | Over past six months, presence and age of onset of various symptoms such as difficulty swallowing, weight loss, excessive sweating, etc. |
| Brain Health Over the Lifecourse | Residential addresses at age 10, 25, 40, 65; physical, cognitive, social activity, weight, and diet at ages 25, 40, and 65; lifelong perceived discrimination (124) (e.g., "A doctor, nurse, or other health care provider might treat me poorly"); coping over the lifecourse (125) (e.g., over lifetime, frequency that participant has felt unable to control important things in life). |

**Table 2.**
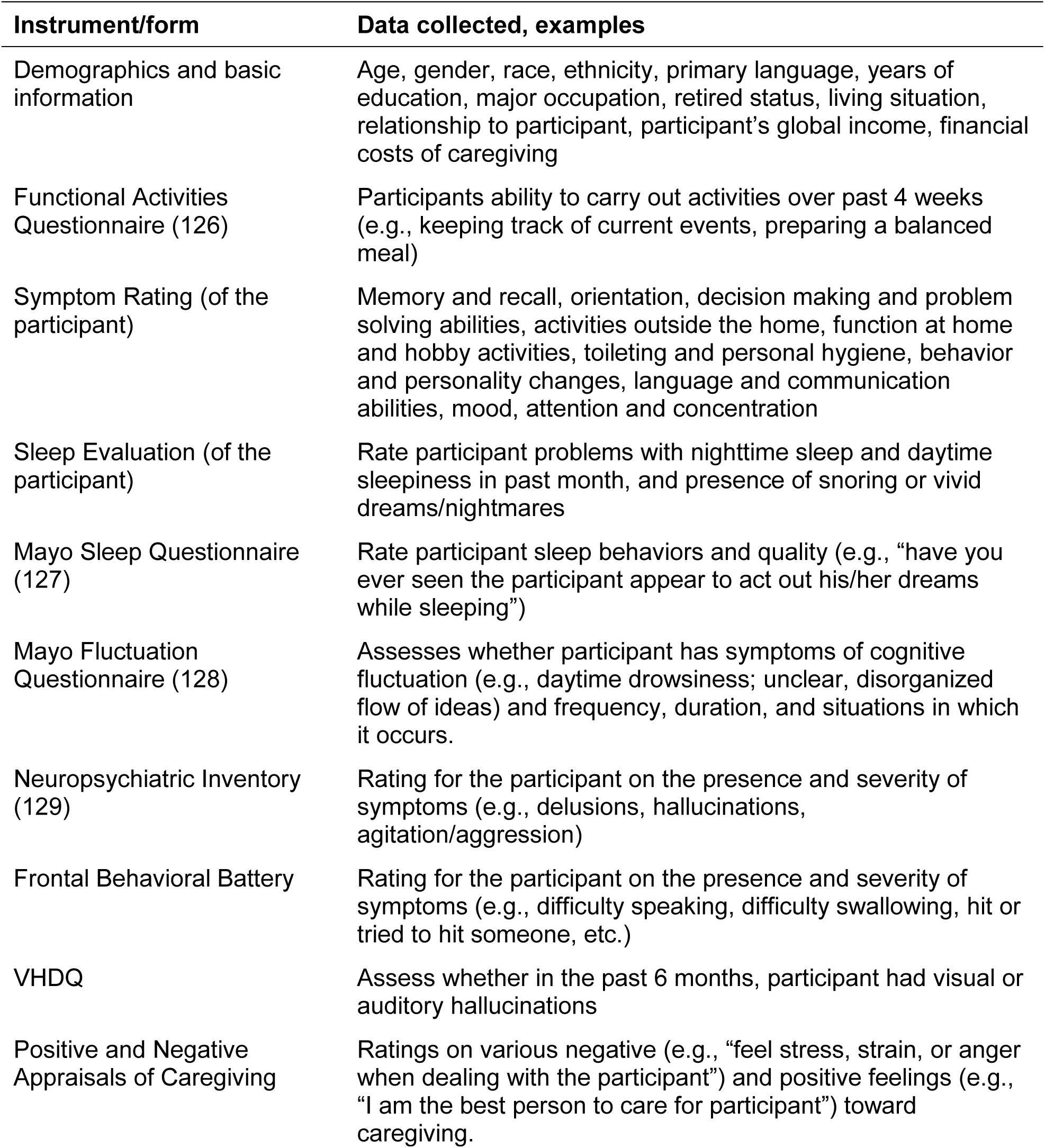
Questionnaires completed by study partner.

| **Instrument/form** | **Data collected, examples** |
| --- | --- |
| Demographics and basic information | Age, gender, race, ethnicity, primary language, years of education, major occupation, retired status, living situation, relationship to participant, participant's global income, financial costs of caregiving |
| Functional Activities Questionnaire (126) | Participants ability to carry out activities over past 4 weeks (e.g., keeping track of current events, preparing a balanced meal) |
| Symptom Rating (of the participant) | Memory and recall, orientation, decision making and problem solving abilities, activities outside the home, function at home and hobby activities, toileting and personal hygiene, behavior and personality changes, language and communication abilities, mood, attention and concentration |
| Sleep Evaluation (of the participant) | Rate participant problems with nighttime sleep and daytime sleepiness in past month, and presence of snoring or vivid dreams/nightmares |
| Mayo Sleep Questionnaire (127) | Rate participant sleep behaviors and quality (e.g., "have you ever seen the participant appear to act out his/her dreams while sleeping") |
| Mayo Fluctuation Questionnaire (128) | Assesses whether participant has symptoms of cognitive fluctuation (e.g., daytime drowsiness; unclear, disorganized flow of ideas) and frequency, duration, and situations in which it occurs. |
| Neuropsychiatric Inventory (129) | Rating for the participant on the presence and severity of symptoms (e.g., delusions, hallucinations, agitation/aggression) |
| Frontal Behavioral Battery | Rating for the participant on the presence and severity of symptoms (e.g., difficulty speaking, difficulty swallowing, hit or tried to hit someone, etc.) |
| VHDQ | Assess whether in the past 6 months, participant had visual or auditory hallucinations |
| Positive and Negative Appraisals of Caregiving | Ratings on various negative (e.g., "feel stress, strain, or anger when dealing with the participant") and positive feelings (e.g., "I am the best person to care for participant") toward caregiving. |

Once the battery of questionnaires is completed and reviewed by the study team for completeness, the baseline visit is scheduled. The visit will include blood sample collection, detailed neuropsychological, physical function, neurological, and psychosocial assessments, and participants receive an actigraphy and peripheral arterial tone device (i.e., Watch PAT) for overnight sleep monitoring and have their MRI scheduled (Table 3). The full baseline visit is expected to take two half days to complete. Participants recruited for certain grant-funded projects that include the HBI assessment also will be scheduled to obtain an amyloid PET using 18F-Fluorbetaben.

**Table 3.** In-person clinician assessments and procedures.

| **Assessment/Procedure** | **Visit done/assessed<sup>a</sup>** |
| --- | --- |
| Blood draw | Baseline visit, every follow-up visit |
| Neuropsychological evaluation | Baseline visit, every follow-up visit |
| Clinical Dementia Rating (CDR®)(32) | Baseline visit, every follow-up visit |
| Physical Function assessment | Baseline visit, every follow-up visit |
| Color Blind Test (45) | Baseline visit, every follow-up visit |
| Physical exam | Baseline visit, every follow-up visit |
| Neurological exam | Baseline visit, every follow-up visit |
| Watch PAT / Sleep study | Baseline visit, every follow-up visit |
| Ankle Brachial Index (ABI) | Baseline visit, every follow-up visit |
| Pulmonary Function Test (PFT) | Baseline visit, every follow-up visit |
| Audiometry Hearing test | Baseline visit, every follow-up visit |
| Optical Coherence Tomography (OCT) | Baseline visit, every follow-up visit |
| Neurobehavioral markers | Baseline visit, every follow-up visit |
| Cognivue (60) | Baseline visit, every follow-up visit |
| Magnetic Resonance Imaging (MRI) | Baseline, every two years afterward |
| Positron Emission Tomography (PET) | Baseline visit (optional) |

<sup>a</sup> IRB-approved modifications to the HBI protocol may add or delete additional assessments/procedures and/or their frequency

The research visit concludes with a feedback session during which a research clinician (e.g., physician, nurse practitioner) meets with the participant and study partner (optional) to go over results of the neurological examination, physical performance, neurobehavioral status, and cognitive evaluation. This meeting also will provide health recommendations, education, resources and referrals based on the findings. This feedback can be shared with family and/or a doctor at the participant’s request. Once reviewed, participants will receive a summary of clinical laboratory results from a CLIA-approved lab. If any abnormalities are detected, the participant will be informed of results and encouraged to speak with their primary care provider for further evaluation. Results of genetic testing will not be shared with either the participant or their physicians as described in the consent form.

#### In-person follow-up visits

Annual follow-up visits will collect almost identical data as the baseline visit, and will include the neuropsychological battery, neurological evaluation, physical performance testing, and blood work, so that we can monitor annual changes in cognitive and physical status and biomarkers for ADRD. MRI are conducted every 2 years following the baseline visit, unless a clinically relevant event (e.g., conversion to dementia, stroke) occurs whereby they may be asked to undergo an MRI earlier.

#### Telephone follow-ups

At 3 and 6 months after the baseline visit and then at 6 months after the follow-up visits, each participant will be contacted by phone to update contact information, health history and medications, collect data on their cognition and everyday functioning, assess adherence with recommendations and referrals provided at the feedback session, and to support study retention.

### Physical and neurological examinations

Each participant will provide information about their current and past health history (e.g., medical, social, physical, environmental) through a 20-page self-administered questionnaire prior to the core visit. To confirm or rule out various types of neurodegenerative diseases (ADRD) a series of tests and examinations will be conducted during the study visit. The results of these procedures will be documented and used to relate objective and subjective findings, measures, and complaints for an accurate research diagnosis for each participant. Each study partner will complete an 11-page survey.

Advanced practice nurses and neurologists will complete a comprehensive physical and neurological examination of each participant (33, 34). This examination will include assessments of vital signs in a sitting and standing position, mental status, cranial nerve, reflexes, sensation, balance, coordination, and motor skills and includes the Unified Parkison’s Disease Rating Scale – Part III Motor Examination (35), modified Hoehn and Yahr staging (36), Lewy Body Composite Risk Score (37), modified Hachinski Ischemic Scale (38), and modified CAIDE scale (39).

#### Overnight Sleep Study

A disposable FDA-approved at home diagnostic test for sleep apnea will be given to each participant to wear overnight following the core visit procedures (40). This actigraphy and peripheral arterial tone device is placed on the wrist of the non-dominant hand, a probe is placed on the index finger of the non-dominant hand, and a sensor is placed on their chest just below the sternal notch. A smartphone is used to accompany the device using an app to start and end the test.

#### Audiometry

Headphones and a tablet computer with auditory testing software will be given to each participant (41). The hearing test will be conducted in a private space free of distractions for a duration of about ten minutes. Participants will be able to use the tablet independently after listening to the instructions provided on the tablet.

#### Spirometry

This test requires the participant to take a deep breath and then expel all the air in their lungs into a filtered mouthpiece that is connected to a spirometry device (42). This machine can measure peak expiratory flow (PEF), PEF%, FEV1 (forced expiratory volume), FEV1%, FVC (forced vital capacity), and FEV1/FVC.

#### Ankle Brachial Index

Cuffs are wrapped around the arm (brachial) and ankle (dorsalis pedis and posterior tibial) arterial sites bilaterally and a handheld doppler ultrasound device is used to measure systolic blood pressure while the participant is in a supine position at rest for 5 to 10 minutes (43). This test will be used to assess peripheral artery disease.

#### Color Blindness Test

The Farnsworth D-15 Dichotomous Color Blindness Test will be administered digitally on a computer or laptop. Participants will be asked to arrange 15 color squares in order by choosing the most similar color and dragging the color squares into the upper boxes (44, 45). A pilot square is set as the first box.

### Cognitive assessments

A comprehensive battery of both traditional (pencil and paper) and computerized tests will be employed to longitudinally assess aspects of cognition and mood. The battery will be conducted at baseline and then repeated at each subsequent annual follow-up. Specific tests and their associated details are provided below.

#### The Clinical Dementia Rating scale

(CDR; 46) is a semi-structured interview, involving both the participant and a study partner, gauges cognitive and functional performance across six domains: Memory, Orientation, Judgment and Problem Solving, Community Affairs, Home and Hobbies, and Personal Care. The CDR has good reliability and validity and detects subtle cognitive changes and everyday functioning. Each domain receives a score between 0 (no impairment) and 3 (severe impairment). An aggregate global score is then derived, indicating overall dementia severity (i.e., 0 =normal, 0.5 = MCI, 1,2,3= mild, moderate, or severe dementia, respectively). The sum of boxes score (CDR-SB) is calculated by summing domain scores with possible scores ranging from 0-18, with higher scores indicating greater impairment.

#### The Global Deterioration Scale

(GDS; (47)) is a rating scale of cognitive status with a range of scores from 1-6. The GDS has advantages over the CDR by providing separate ratings for cognitively normal individuals with and without cognitive complaints. GDS 1 signifies cognitively normal individuals without any subjective complaints; GDS 2 signifies cognitively normal individuals with subjective complaints; GDS 3 signifies the earliest objective cognitive deficits equivalent to MCI; and GDS 4 signifies clear cut cognitive and functional deficits equivalent to mild dementia. GDS 5-7 designate moderate through severe stages of dementia.

#### The Hopkins Verbal Learning Test-Revised

(HVLT; 48) is a neuropsychological assessment tool used to assess verbal learning and memory. Participants are instructed to recall a list of 12 noun-words over three trials. Total words correctly recalled across three trials (max 36). The number of correctly recalled words following a 20–25-minute delay (range 0-12) is used as a measure of free recall. The number of correctly identified words from the yes/no recognition task (range 0-12) is used as a measure of cue recognition memory.

#### The Montreal Cognitive Assessment

(MoCA (49)) is a widely utilized cognitive screening instrument designed to identify mild cognitive impairment. Cognitive domains assessed include Visuospatial/executive functions, language, memory attention, delayed recall and orientation. The MoCA has a total possible score of 30, with a score of 26 or above considered within a normal range, while scores between 18 and 25 are suggestive of MCI, and below 18 is suggestive of dementia.

#### Number span – Forwards

(50) is used as a measure of simple auditory attention. Participants are presented with an increasing sequence of numbers over several trials and are required to repeat them back. The longest sequence correctly repeated is used as the measure of attentional capacity with a score range of 0-9.

#### Number span – Backwards

is used as a measure of auditory working memory. Participants are asked to repeat number sequences in reverse order. The longest backward-repeated sequence is used as the measure of working memory with scores ranging from 0-8. For forwards and backwards span, higher scores denote better performance.

#### Noise Pareidolia Task - Short Form

(51) is a neuropsychological assessment designed to evaluate an individual’s propensity to perceive faces within ambiguous black and white patterns, offering insights into the phenomena of pareidolia and potential neurological or psychiatric implications of this perceptual bias. Following a practice trail, participants view multiple patterned items. For each, they respond to, “Do you see a face?”, marking the perceived face’s location if positive. Scoring categories include Correct Face Responses with a range of 0-7, indicating accurate identification of actual faces; Correct Noise Responses ranging from 0-13, denoting accurate rejection of patterns without faces; a Total Correct score combining the two previous scores with a range of 0-20; and Noise (Pareidolia) Responses, capturing incorrect identifications of faces in noise-only patterns, with a possible range of 0-13. A high score indicates better perceptual discrimination, while frequent false positives (seeing faces where none exist) might point to a heightened propensity for pareidolia.

#### The Animal Naming task

(52): This is used is a widely used measure to assess semantic fluency. Participants are asked to name as many different animals as quickly as possible within 60 seconds. The total score represents the number of distinct animals named within the allowed time, and higher scores indicate better semantic fluency and retrieval ability.

#### The 32-item Multilingual Naming Test

(MINT; 53) is a standardized instrument designed to quantify confrontation naming ability. Participants are presented with a set of 32 pictorial stimuli, each representing distinct objects, and are tasked to elicit the corresponding lexical representation. The MINT aims to assess lexical retrieval and object recognition capacities, integral elements of linguistic processing. Furthermore, the test’s construct is curated to mitigate biases across diverse linguistic and cultural demographics. Scoring is systematic: each correctly identified stimulus contributes one point, culminating in a maximum possible score of 32, which is indicative of optimal linguistic processing capability.

#### The Trail Making Test

(TMT; 54) is a widely recognized neuropsychological assessment used to examine cognitive flexibility, visual search, motor speed, and executive functioning. Comprising two main components, Trails A and B, the test evaluates both simple psychomotor speed and more complex task-switching abilities. <u>Trails A</u> requires participants to connect numbered circles (1–25) in ascending order as quickly as possible. The time taken in seconds to complete is used as a measure of processing speed. The max time to complete the test is 180 seconds. Longer completion times suggest poorer performances. <u>Trails B</u> is a widely used measure of task switching, cognitive flexibility, and executive functioning. In this task, participants are asked to connect numbers and letters in ascending order (e.g., 1-A-2-B-3-C, etc.). The completion time in seconds is used as the outcome measure. The max time to complete the test is 300 seconds.

### Longer completion times suggest poorer performances

#### Number symbol coding

(55): The NSCT is a brief measure of processing speed, attention, and working memory that can be completed by individuals into the moderate-to-severe stages of dementia. In the task, participants are typically required to match numbers with specific symbols within 90 seconds. Its streamlined nature allows for efficient administration. A score of 36 or below indicates cognitive impairment with possible scores ranging from 0-70.

#### The AD8

(56) is an informant-based interview designed to assess for signs and symptoms of dementia. The instrument is grounded in the principle that study partners (often close family members or friends) can provide crucial insights into an individual’s cognitive and functional changes over time. The AD8 evaluates various domains, including memory, orientation, executive function, and interest in activities, through a series of eight questions. The study partner is asked to compare the individual’s current cognitive and functional performance with their performance in the past. Responses indicating change consistent with cognitive decline are scored, with a higher score suggesting a greater likelihood of dementia. A score of 2 or more on the AD8 indicates that further dementia evaluation is warranted.

#### The Clock Drawing Test

(CDT (57)) is an established neuropsychological screening tool used to assess visuospatial abilities, executive functioning, and attention. This test requires participants to draw the face of a clock on a blank sheet of paper and then set the hands to a specified time, “10 minutes past 11.” Scoring the CDT can range from a basic dichotomous “normal/abnormal” rating system that consider various aspects of the drawing, such as the presence and correct sequencing of numbers, correct placement and proportion of the clock hands, and overall spatial organization.

#### The Craft Story

(58) is a measure of verbal episodic memory. It consists of a short narrative that is read aloud to participants, who are then asked to recall as much information from the story as possible immediately after hearing it (Immediate Recall) and again after a specified delay (Delayed Recall), approximately 20 to 30 minutes later. Scoring is typically based on the number of thematic units (score range: 0-25) or specific details accurately recalled verbatim (score range: 0-44). Higher scores indicate better verbal learning and memory.

#### The Benson Complex Figure Test

(BCFT; 59) is used to assess visuoconstructive skills, visual memory, and recognition. Participants begin by drawing a copy of a complex geometric figure (“copy” trial). After a delay of approximately 30 minutes, they are asked to redraw the figure from memory (“delayed recall” trial). Lastly, in the “recognition” trial, participants are asked if they recognize the figure from an array of figures with a yes-no response. Scoring is based on the accuracy and organization of drawings, types of errors in reproduction, and the accuracy of recognition. The BCFT is particularly useful for identifying cognitive deficits, especially those associated with right hemisphere dysfunctions. For the copy and delayed recall trial, a possible score of 0-17 can be obtained, with higher scores indicating better performances.

#### Cognivue®

(60) is a computerized comprehensive cognitive assessment tool that evaluates a range of specific cognitive domains, including motor speed, visuospatial abilities, verbal and visual episodic memory, attention, executive functioning, and reaction time. Using adaptive psychophysics, it adjusts task difficulty in real-time based on participant responses. The assessment, which takes about 10 minutes, provides a numerical score categorizing performance as “normal,” “mild impairment,” or “moderate impairment.” Cognivue® is Food and Drug Administration (FDA)-cleared for use as an adjunctive tool to aid in assessing cognitive impairment in participants aged 55-95 years.

### Anxiety and depressive symptoms

#### The Hospital Anxiety and Depression Scale

(HADS; 61) is a validated instrument for screening symptoms of anxiety and depression in medically ill populations, specifically within non-psychiatric hospital contexts. The HADS encompasses 14 items, divided into two subscales: the Anxiety subscale (HADS-A) and the Depression subscale (HADS-D). Each item is appraised on a 4-point Likert scale, resulting in individual subscale scores that can range from 0 to 21.

Interpretation of scores on both subscales is as follows: 0-7: Normal; 8-10: Borderline; 11-21: Abnormal. The HADS is recognized for its strong psychometric properties and has been employed extensively across various clinical cohorts.

#### Depression, Anxiety, and Apathy Assessment (DA^3^)

is a 21-item questionnaire assessing how participants have felt and their experiences during the previous 2 weeks. Questions such as “I feel tense and unable to relax”, “I am able to get things started and completed”, and “I have trouble focusing my thoughts” are answered on a 4-point scale (0=rarely, 1=sometimes, 2=often, or 3=always). Items are summed to provide a depression score (DA3-D, range: 0-7), an anxiety score (DA3-An, range: 0-7), and an apathy score (DA3-Ap, range: 0-7).

### Physical function assessment

At baseline and each annual follow-up visit, HBI participants will undergo extensive performance-based physical functional assessments including body composition, muscle strength, balance, physical performance, and computerized gait analysis. Participants will be asked to come in wearing comfortable clothing and shoes and will start the physical functional assessment with a series of body measurements (i.e., height, hip-, waist-, and neck-girth) taken with wall-mounted or handheld tape measures. Weight will be measured with InBody, a bioimpedance-based body composition analyzer, which provides estimates of weight, BMI, skeletal muscle mass, fat mass including visceral fat, intracellular and extracellular water content, basal metabolic rate, and other body composition indicators (InBody USA; 770 series).

Muscle strength will be measured in the upper extremities with a Smedley Digital Hand Dynamometer (Baseline® Evaluation Instruments, Version 12-0286) for a total of three trials in each hand, with the average across the three trials used in data analysis. *Sarcopenia* status is reported using the Short Portable Sarcopenia Measure (SPSM) (62). The SPSM score ranges from 0-18, with lower scores indicating greater likelihood of sarcopenia.

The mini-Physical Performance Task (mPPT) consists of 4 tasks: (1) Bending Over Test for which participants will be asked to bend over without bending their knees to pick up a binder clip placed about 1 foot from their foot on the dominant side; (2) a 50-foot timed walk task, (3) the 5 Times Sit to Stand Test (5XSST) using a sturdy 4-leg non-rolling chair with hand support, with the participants asked to rise and sit down in the chair as fast as they can for a total of five times without using the armrests; and (4) the Progressive Romberg Balance Test (PRBT) for which participants are timed as they complete 10-second side-by-side, semi-tandem, and tandem balance tasks (63). The mPPT score ranges from 0-16 with higher scores indicating greater physical functionality.

Participants will undergo a complex gait and balance evaluation on an electronic pressure-based walkway (Zeno Walkway^TM^) and data processed using the ProtoKinetics Movement Analysis Software (ProtoKinetics, Version 6.00C1). The Zeno Walkway provides a wealth of temporal and spatial measures of gait associated with fall risk including velocity, cycle time, % stance, step length, stride width, % swing, % single support, and other kinetic and kinematic gait parameters. The HBI protocol requires completion of a series of walking tests performed on the Zeno Walkway in the following sequence: (1) Single-task (walking at normal pace + no concurrent cognitive task); (2) Dual-task (walking + reciting the alphabet); (3) Dual-task (+ serial subtraction); and (4) Dual-task (+ texting) with parameter estimates derived for each of the four walks.

Balance and risk of fall also will be assessed with 1) Limits of Stability (LOS) and 2) Four Square Step Test (FSST). LOS measures how much an individual can lean in four directions: the mediolateral (left/right) and anteroposterior (forward/backward) directions/axes while standing with their feet shoulder width apart. LOS starts with the participant standing for 30 seconds in quiet standing (QST) with their arms by their side with eyes open. Participants are then asked to lean forward as far as they can without taking a recovery step or falling, return to center, then lean left, return to center, lean backward, return to center, and finally lean right and return to center. While no specific amount of time holding the lean is required, the rater is assessing whether the lean is held for a minimum of 2 seconds. One trial is conducted for the LOS and QST tests, with one retake allowed. FSST is a dynamic test of balance and coordination which involves stepping as fast as possible in each of 4 squares with both feet in a predetermined sequence (64). Participants are allowed to practice one time before testing commences with one retake permitted in case of tripping or incorrect step sequence. Two trials are completed for FSST, with the better time (in seconds) being recorded as the score. Both LOS and FSST are conducted with the Zeno Walkway inertial sensor waist belt to capture trunk excursion.

Timed up and Go (TUG) is a timed assessment that requires a person to stand (handsfree) from a chair walk 3 meters and return to sitting (handsfree) (65, 66). It is considered a gold standard indicating falls risk in older adults. Cognitive TUG i(TUG-Cog) is TUG with the addition of a cognitive task such as serial subtraction (e.g., subtract 87 by 3).

Kinetisense is a markerless motion capture analysis system that uses infrared cameras to identify the kinematics of moving joints (67, 68). It provides real-time biofeedback on the performance of balance, gait, and functional activities in coronal, sagittal, and transverse planes. Kinetisense produces video playback and generated reports that include a risk of fall gait assessment that is calculated based on normative gait parameters and when combined with other gold standard physical performance assessments such as the TUG and STSX5, provides ergonomic feedback of a person’s overall function and balance.

Three global measures of frailty are determined at each visit. The Rockwood Frailty Index (69), the Fried Frailty Phenotype (70), and the CHSA Clinical Frailty Scale (71). Higher scores on the frailty scales are suggestive of greater likelihood of being frail.

### Blood- and saliva-based biomarkers and genetics

A custom panel of tests (i.e., Healthy Brain Initiative (HBI) Profile) will be conducted. At the start of the core visit, blood will be drawn from each participant by venipuncture by trained phlebotomists. Participants also provide saliva samples for DNA extraction and APOE e4 carrier status. Saliva will be obtained through self-collection kits. The collected specimens will be processed and used to evaluate plasma biomarkers, genetic data, and metabolic panels, and then stored in a biorepository for future research as plasma, whole blood, and DNA. Measures collected include a comprehensive metabolic panel, cortisol, cystatin C, hemoglobin A1C, insulin resistance, homocysteine, high-sensitivity C-reactive protein (hs-CRP), a complete lipid panel with particles, thyroid stimulating hormone, vitamin B12, folate, vitamin D, apolipoprotein E (APOE) e4 status, and multiple plasma ADRD biomarkers (e.g., Ab42, Ab40, p-tau). Blood specimens will be processed in HBI’s lab for shipment to C2N laboratories for analysis. C2N employs mass spectrometry to calculate multiple plasma AD biomarker values including Ab42/40, p-tau181, p-tau217, and p-tau/np-tau. Additionally, the HBI lab conducts in-house measurements of biomarkers such as neurofilament light (NFL), glial fibrillary acidic protein (GFAP), and TAR DNA-binding protein 43 (TDP-43) on Quanterix SIMOA™ platforms for each participant.

### Machine-based assessments

*Optical coherences tomography* (OCT), an imaging technique using near-infrared light to capture detailed images of the cornea, lens, retinal tissue, anterior chamber, and retinal vasculature, is an emerging technology in ADRD research. The retina is embryologically derived from the same tissue as the cerebral cortex, and pathological changes in cortical gray matter appear in the retina as well, including deposits of amyloid-β revealed using targeted tracers (72, 73). Retinal measurements include the central subfield thickness, the volume and thickness of the macula, the thickness of the retinal nerve fiber layer, internal plexiform layer thickness, and the ganglion cell layer thickness. The minimum thickness of both the ganglion cell layer and the internal plexiform layer is also collected and is strongly associated with various cognitive domains and the CDR sum of boxes along with the combined ganglion cell layer and internal plexiform layer thicknesses (74). By collecting these data in HBI, we aim to explore the use of these markers in determining current impairment through non-invasive imaging procedures as well as identifying risk of future impairment based on the health of retinal tissue (75–77).

*Neurobehavioral markers*, including gaze behavior and speech behavior, have been shown to identify longitudinal changes in prodromal dementia, with some research showing an ability to identify preclinical impairment through the examination of connected speech production (78–80). Gaze behavior assessed through eye movements during cognitive assessment tasks further capture differences between not only cognitively normal participants and impaired participants through the examination of exploratory behavior and novelty preference, but differentiation of likely cause of impairment including separating DLB and PD from AD using saccadic behavior (75, 81, 82). Eye movements are collected using Tobii Pro eye trackers, including the Tobii Dynavox and the Tobii Pro Spectrum, and audio metrics including speech are collected using hypercardioid directional microphones for reducing extraneous background noise. These markers are collected while picture description, narrative recall, and visual search tasks are administered to participants using custom-designed experimental software written in Python and C++.

### Neuroimaging procedures and measures

#### MRI

All participants will have brain MRI scans using a GE 3 Tesla 750W scanner at baseline and every 2 years. We will collect high-resolution 3D sagittal Magnetization-Prepared Rapid Gradient-Echo (MPRAGE) images with axial and coronal reconstructions to quantitate brain atrophy. We will use axial 3DT2 FLAIR and 2D T2* brain images to quantify white matter hyperintensities (WMH), infarcts, microhemorrhages and superficial siderosis. We will collect diffusion-weighted images (diffusion tensor imaging) to measure mean diffusivity and fractional anisotropy to estimate microstructural integrity and white matter tractography. Last, we will collect resting BOLD sequences for functional MRI analyses. The total imaging time sequences is estimated to be 45 minutes. The MRI sequences selected for this project are modified from the ADNI-3 canonical protocol and were selected to enhance compatibility with national efforts (83).

In addition to visual reads for MRI scans, post-processing of all MRI images will be performed according to ADNI procedures, including correction for geometric distortion of gradient nonlinearity, field inhomogeneity correction, histogram-peak sharpening, and correction for site-specific scanner features/field strength. Cortical, subcortical, ventricular, and white matter hyperintensity volumes are analyzed using Combinatics® cNeuro Suite, an FDA-cleared quantitative pipeline. Automated brain imaging quantification using artificial intelligence (AI) technology offers many opportunities to reduce the time needed for imaging analysis and increase the accuracy, precision, consistency, robustness, and, as a consequence, the research quality of the data. Combinostics cMRI suite is comparable to manual segmentation with Pearson coefficients ranging from 0.84 to 0.99. The cMRI suite contains important dementia biomarkers including hippocampal, inferior lateral ventricle, cerebral white and gray matter, cerebral cortex, and medial temporal lobe volumes, and a global cortical atrophy index with an 85% sensitivity and 86% specificity. White matter integrity and myeline measures will be conducted using Synthetic MRI®, an FDA-cleared AI pipeline providing quantitative measurements of intracranial, brain parenchyma, myelin, cerebrospinal fluid volumes and brain parenchymal and myelin correlated fractions.

#### PET

Participants who consent will have baseline amyloid PET scans with ligands from Life Molecular Imaging. Non-contrast enhanced computerized tomography (CT) is used for attenuation correction of the PET images and for morphological localization through image fusion between PET and CT. PET scans will be processed using the reconstruction algorithm provided to include attenuation correction with an image matrix size of 128 x 128, and a transaxial pixel size of 2-3 mm. Participants will undergo an amyloid PET scan using Neuraceq™ (F18 Florbetaben injection), an amyloid PET ligand that received FDA approval in March 2014.

8.1 mCi +/-20% of Neuraceq will be administered as a single bolus intravenous injection at a rate of 6 seconds/mL in a total volume of up to 10 mL. Approximately 10 minutes prior to the initiation of the PET scan, the participant will be positioned in the PET scanner. The head will be positioned such that the entire brain including the cerebellum will be in the field of view (FOV).

An attenuation correction image will be acquired following standard procedures. PET imaging will commence 90 min post injection for a total of 20 min (4 x 5 min frames). SUVR images will be created using 90 min after injection and used for qualitative reads and to extract global values. The Combinostics cPET® suite provides a segmental analysis of FDG and amyloid PET with regional and global SUVRs and for amyloid PET calculates the Centiloid Level.

### Data and biospecimen repository

The data repository will consist of clinical, cognitive, behavioral, and functional data as well as summary data from plasma, DNA, and imaging biomarker studies that will be kept in both paper and electronic form and kept in locked cabinets within locked offices and compiled in a REDCap electronic database accessible via protected passwords. The database will contain all data derived from interviews of participants and study partners, neuropsychological and physical performance assessments, medical exams, as well as summaries of blood and imaging data. Physical records of neuroimaging studies will be placed in participant files kept in locked file cabinets and summary results will be kept in a separate protected electronic file.

Blood specimens are processed in the lab for shipping to commercial laboratories for analysis and for storage in -80°C freezers located in the lab (with generator backup). Buffy coat will be extracted from blood for future DNA studies. All specimens collected will be stored in the CCBH laboratory refrigerator and freezers, which are only accessible by CCBH laboratory personnel. The specimens will be kept indefinitely.

Biological samples will be sent to the National Centralized Repository for Alzheimer’s Disease and Related Dementias (NCRAD) for sharing with other investigators. NCRAD is a national resource supported by the National Institute on Aging (NIA) that prepares and stores biological specimens from all over the world and makes them available to approved scientists who would not otherwise be able to access this material. Biological samples will be de-identified and maintained in the CCBH laboratory indefinitely. These biological samples allow for studies of the causes of many diseases, including ADRD.

### Primary HBI outcomes

The primary outcome of HBI is to improve our understanding of what healthy brain aging is, and how to best detect the earliest signs of MCI/ADRD in diverse populations. This is accomplished via deep phenotyping, biomarker collection, and genetic (including epigenetic) markers. This work has allowed us to develop the brain health platform (described below) and leverage the ATN framework to begin to take a precision medicine approach to brain health and ADRD detection, and eventually treatment and prevention.

#### Brain health platform

The Brain Health Platform (84) quantifies brain health and identifies ADRD risk factors based on 3 measures collected that will be collected at HBI: Resilience Index (RI) (85), the Vulnerability Index (VI) (86); and the Number-Symbol Coding Task (NSCT) (55). VI and RI scores (range: 2-20 and 1-378, respectively) are calculated from demographics, lifestyle questionnaires, physical assessments, medical history, and neuropsychological examination. Details of the Brain Health Platform including the VI and RI are published elsewhere (85, 86). By combining measures of performance, resilience, and vulnerability, it provides an overall snapshot of the participant’s brain health at a given visit. Instead of a summary score, the platform indicates whether participants are at low, intermediate, or high risk of being cognitively impaired.

#### ATN Staging

Once a sufficient number of individuals have completed MRI, amyloid and tau PET, and plasma ADRD biomarkers, we will use the distribution of values within the normal control group to z-score individual participants and provide categorization consistent with the ATN (amyloid tau neurodegeneration) research framework (13). Given the bimodal and global distribution of amyloid PET data, Gaussian mixture modeling will be used to identify A- and A+ distributions. Categorization of Tau PET into Braak III and above will be used to classify individuals as T- or T+ and hippocampus volume will be used to classify individuals as N- or N+. This will be matched to plasma Ab40, Ab42 (A- and A+), Tau and p-tau (T- or T+), and NFL (N- or N+). Further, we can consider other pathologies in the form of MRI white matter hyperintensities for vascular disease (V- or V+), plasma alpha synuclein and phospho-synuclein for Lewy body pathology (S- or S+), and plasma TDP-43 (P- or P+). This will permit testing of the ATN framework in a systematic fashion in a deeply phenotyped cohort.

#### Clinical diagnoses

A clinical diagnosis of the participant’s cognitive status (normal cognition, MCI, or dementia) and their ADRD etiological diagnosis (e.g., Alzheimer’s disease or Lewy body disease) is determined via consensus conference of the HBI clinicians. To make the diagnosis, clinicians review the neuropsychological test findings and supporting clinical data independent of the biomarker data and follow consensus criteria guidelines (2, 3, 87–90).

### Intermediary/secondary outcomes

Beyond the ATN framework, Brain Health Platform, and clinical diagnoses, which capture presence of disease or risk of developing disease, continuous measures of the ADRD biomarkers and functional and cognitive test scores (e.g., longitudinal change in plasma Ab42/40, frailty scores, or executive function over time) will serve as intermediary outcomes.

### Novel approaches in the HBI

#### Precision medicine

Precision medicine is a model that permits the separation of patients into different groups to better inform and tailor medical decisions, practice, and treatment to defined subgroups based on their (a) predicted responses and/or (b) risk of disease (91–93). Precision medicine approaches can help shift medical practice from reaction to prevention, predict susceptibility, improve early detection, and eliminate trial and error treatment thereby avoiding or limited negative side effects and maximizing efficacy (94). Commonly used in oncology, precision medicine has expanded to other fields. Precision medicine approaches are typically genome-based using tissue markers acquired via biopsy or genetic markers in blood. In the case of ADRD, biopsy is not a viable path and genetic causes of disease are limited. Instead, HBI has focused on deep phenotyping individuals to examine small deviations from normal or expected structure, function, and behavior. Using this detailed and comprehensive picture, HBI will better characterize factors that make individuals (a) more likely to develop disease (i.e., vulnerability) or less likely to develop disease (i.e., resilience) (95, 96).

#### Machine learning

Given the vast quantities of data generated through the HBI, machine learning techniques become viable and, in some cases, preferable compared to more traditional statistical techniques of data analysis. Model generation using supervised learning techniques, including both classification and regression, enables hypothesis testing of diagnostic and screening techniques, leading to developments such as multi-stage screening systems and the identification of markers associated with early-stage and prodromal cognitive impairment including groups of cognitive assessments as well as markers of vulnerability and brain health (84, 86, 97, 98).

#### Geospatial data and the exposome

HBI will collect residential addresses and self-reported environmental exposures and behaviors from various time points during the participant’s life course from childhood to later life. These data will address HBI’s specific aim to determine how sociodemographic and lifestyle factors contribute to risk for and resilience against ADRD. HBI’s life course measures will target key domains of the exposome, which has been defined as the “exposures of an individual in a lifetime and how those exposures relate to health” and encompasses all non-genetic determinants of health (99). Domains to be captured in HBI center on “general external” exposome measures of social context (e.g., participant’s and neighborhood SES) and the neighborhood built environment (e.g., greenspaces, sidewalk availability), as well as “specific external” exposome measures encompassing lifestyle behaviors (e.g., diet, exercise, medication use). The measures will either be collected via self-report by the participant or study partner or will be derived by geocoding addresses to identify the latitude-longitude and/or Census tract location and linking to built and social environment data from sources such as the US Census, the Neighborhood Atlas (100), and satellite imagery from the US Geological Survey (101).

#### Epigenetics

DNA methylation (DNAm) is an epigenetic modification where a methyl group attaches to DNA’s cytosine nucleotides, often at CpG sites (102). This process can vary with cell type, developmental stage, and environmental conditions (103, 104) has emerged as a pivotal biomarker of biological aging, with its profiles often referred to as “epigenetic clocks” (105).

These clocks hint at disparities between chronological and biological age, with greater than expected biological ages associated with increased risk for neurodegenerative disease and earlier mortality (105–107). Specific lifestyle risks, such as hypertension (108, 109) and obesity (110), correlate with distinct DNAm changes in genes governing vascular function, neuroinflammation, and cognition (111, 112). In essence, DNAm may offer a potential pathway through which cognitive health disparities may arise.

Our objective for collecting DNAm data will be to improve and refine current epigenetic age clocks to brain aging while also seeking an in-depth investigation of associations between DNAm profiles and various lifestyle factors. This endeavor is geared towards a comprehensive understanding of the relationship between DNAm alterations and brain health, with a focus on the molecular determinants underpinning aging processes. Whole blood samples (10 mL) from HBI participants will be used for DNAm data extraction. DNAm extraction will be performed using the Infinium Methylation EPIC BeadChip 850k platform the Center for Genome Technology at John P. Hussman Institute for Human Genomics, University of Miami Miller School of Medicine. Using these data, our objective is to construct refined epigenetic clocks that provide enhanced precision in estimating biological age and its divergence from chronological age.

#### Transdisciplinary and multidisciplinary study team approach

The HBI team was purposefully constructed to be multidisciplinary and transdisciplinary, including individuals from varied disciplines who continuously collaborate, infusing conceptual knowledge and methodologies unique to each discipline to inform not only the cohort study protocol but also in the methods, analyses, and means of reporting results when addressing the HBI’s specific aims. The multidisciplinary team includes physicians, nurse practitioners, nurses, social workers, physical therapists, gerontologists, cognitive neuroscientists, biostatisticians, bioinformatic analysts, molecular scientists, data scientists, geographic information system specialists, and an urban planner. The HBI team structure and personnel expertise/skills will be modified as needed to increase efficiency and accommodate new research initiatives.

### Data management plans and data security

Data management is handled by the CCBH Data Core, which includes a Data Manager, Data Scientists, and trained data entry personnel. CCBH uses a multimodal data collection approach that includes both paper (e.g., neuropsychological battery) and electronic sources (e.g., participant and study partner questionnaires). Paper sources are scanned and entered in REDCap, an electronic data capture and management system offered through the University of Miami Miller School of Medicine. Access to REDCap via password and restricted to CCBH Data Core staff. Participant and study partner questionnaires are distributed via email to individual study participants and accessed via REDCap-generated questionnaire link, with completed questionnaires saved directly in REDCap. Select parameters from machine (e.g., OCT, Zeno Walkway) and lab and imaging (e.g., MRI, PET) reports are entered in REDCap with full reports stored on University of Miami approved and backed-up shared drives.

REDCap is managed by the University of Miami Biomedical and the Health Research Informatics Core, a secure, private cloud and isolated network infrastructure. This isolated health network is protected by an independent firewall; remote access is provided by a Virtual Private Network (VPN). Data security is assured by Virtual Routing Forwarding (VRF); 2Factor VPN authentication; 10 Gbps Juniper Firewall; Web Proxy Security; Load Balancer; Dell XC Web-Scale Converged Appliances running Windows Hyper-v with Azure; Desktop Virtualization with Desktop connections; and Disaster Recovery. The VRF is an isolated, protected network perimeter that enforces the necessary security and compliance restrictions needed to protect data from external threats.

All precautions to protect the integrity of the data are taken and any unauthorized access will be reported to the University of Miami Institutional Review Board. To minimize risk, all materials are kept in locked file cabinets (paper sources) and secure servers (electronic sources), which are only available to the research team members. Data shared outside of the study team is de-identified prior to distribution using a unique participant code identifier. The CCBH laboratory has restricted access to personnel. Samples collected are processed immediately and sent out for analysis. Excess blood is stored in the CCBH laboratory refrigerator and freezers indefinitely, according to current standard operating procedures.

### Safety considerations

This observational cohort study does not meet the definition of a clinical trial and an external Data Safety Monitoring Board is not required. Despite all precautions, medical complications may occur and any adverse events will be reported to the IRB. These include not only physical injury, but also possible psychological, social or economic harm, discomfort, inconvenience, or breach of confidentiality. To minimize risk, all study materials are kept in locked file cabinets and only available to investigators and research personnel associated with the study. In the event of an adverse effect, medical personnel are available with access to an emergency room. For the standard clinical examinations, there are minimal to no risks associated with testing. The main risk is loss of privacy, and every precaution is taken to ensure confidentiality. We are sensitive to disclosure of examination results; this will be performed in private with feedback including medical referrals and follow-up provided to participants. All precautions to protect the integrity of the data repository and registry will be made and any unauthorized access will be reported to the University of Miami Miller School of Medicine IRB.

Following the feedback session, participants can be referred to CCBH clinical practices for clinical evaluation and follow-up. There are no inherent risks associated with the physical, neurological, or psychiatric exams other than discovering a condition that was not previously known. Specific HBI procedures have specific risks as described below:

- To reduce the risk of loss of balance and falling, functional testing will be performed by research staff trained by a physical therapist with advanced expertise (DPT). There is a potential risk in self-collected saliva, of coming into contact with the stabilizing fluid located in the test tube. Users may experience eye and/or skin irritation. User instructions provide warning and how to obtain guidance for handling accidental contact with the stabilizing fluid. The potential risks of blood drawing may occasionally include pain, bruising, fainting, or a small infection at the puncture site. Indications of procedure to follow in case of any of these will be provided to the participants.
- MRI scanning involves the use of a magnet and radio frequency energy. Individuals who have implanted devices, such as pacemakers, certain aneurysm clips, shrapnel, defibrillators, cochlear/ear devices, deep brain stimulators, spinal cord monitoring devices, or metal in the eye or skull are at risk. Therefore, before the procedure, each candidate will be pre-screened for presence of absolute and relative contraindications. Presence of absolute contraindications will disqualify participants from the MRI procedure, while presence of relative contraindications will warrant a careful weighting of risks against benefits for the participant. Those cleared for the procedure will be instructed to remove all metallic objects prior to entering the MRI room. In the case of a medical emergency, trained personnel will follow their respective MRI Emergency Procedures Protocol, which entails calling 911, transferring the participant out of the magnetic field area, and providing basic life support by the radiologist, if necessary, until emergency service arrives. Procedures are also described for situations when an emergency stop is required, a magnet emergency occurs, or other unexpected events (e.g., power outages) emerge.
- The risks associated with PET scans will be discussed with the participants prior to them undertaking the procedure. They include exposure to a radioactive ligand and local site injection reactions. The most common adverse reactions are injection site erythema (1.7%), injection site irritation (1.1%) and injection site pain (3.4%). It is not known whether Neuraceq can cause fetal harm (Pregnancy Category C) so women of reproductive potential will be required to have a pregnancy test (paid for by the study) to assess pregnancy status prior to administration. Pregnant women are excluded from participating in this component of the study. PET compounds, such as Florbetaben F18 (Neuraceq), use small amounts of radiation to generate images. Everyone receives a small amount of unavoidable radiation each year. Some of this radiation comes from space and some from naturally occurring radioactive forms of water and minerals. Each Florbetaben F18 scan has a radiation dose equivalent to about 1.6 years’ worth of natural radiation (approximately 5.8 millisievert or mSv). The use of a CT scan to calculate attenuation correction for reconstruction of Neuraceq images (as done in PET/CT imaging) will add radiation exposure. Diagnostic head CT scans using helical scanners administer an average of 2.2 ± 1.3 mSv effective dose (CRCPD Publication E-07-2, 2007). The actual radiation dose is operator and scanner dependent. Thus, the total combined radiation exposure from Neuraceq administration and subsequent scan on a PET/CT scanner is estimated to be 8 mSv.

### Sample size/power and planned statistical analyses

#### Sample size and power

HBI encompasses multiple aims (described further above), and therefore varying statistical approaches and sample sizes will be involved depending on the research question at hand. Diagnostics will be conducted to assess for non-linearity, missing data, and other issues (e.g., multicollinearity) and adjustments will be made to the analyses as necessary (e.g., multiple imputation).

The HBI was originally built off and thus devised to accommodate planned analyses for a grant-funded study on Multicultural Community Dementia Screening” (MCS) (NIH/NIA R01AG040211). The MCS study was designed to evaluate transition in cognitive status according to NIA-AA stages. For that study, we calculated that we would need a sample of 500 participants to give us a representative sample across racial and ethnic strata, sex, and NIA-AA stages I-IV. This works out to approximately 100 individuals for each of the 5 cognitive states (e.g., normal cognition, subjective cognitive decline, etc.). To maintain 80% power for Black participants based on the desired racial/ethnic distribution (80% White, 20% Black; 35% Hispanic), we will need at least 100 (20% with 95% C.I. 82 - 118) for a minimum prevalence rate of 28%.

#### Statistical analyses

The MCS study aims to detect longitudinal decline and transition across NIA-AA stages using the baseline measures and develop a predictive model for identifying individuals at high risk to transition between NIA-AA stages. Several statistical approaches will be used to characterize longitudinal changes within a multicultural community-based cohort to determine whether baseline demographic, functional, genetic, and medical characteristics predict changes in cognition from baseline during annual evaluations. The progression of prevalence rate changes from baseline to 12, 24, and 36 months of follow-up will be calculated with 95% CI using Clopper-Pearson interval (113). During planned follow-up, we estimate that approximately 5% of cognitively normal participants aged 55-65, 10% of cognitively normal participants aged 65-75, and 40% of cognitively normal participants aged 75+ at baseline will transition to some form of cognitive impairment (114). We conservatively estimate a 15% attrition rate based on published rates (115) – this is higher than our experience with prior cohorts (∼10%).

For longitudinal cognitive decline and transition across NIA-AA stages, we will use generalized linear mixed effect (GLME) models to compare the baseline diagnostic groups on rate of decline in cognitive performance. GMLE models also allow the assessment of change in continuous outcomes measure at multiple time points, modeling both fixed and random effects, and flexibility handling missing data. The impact of baseline cognitive status on the cumulative odds of being cognitively impaired (SCI, MCI and ADRD) versus being cognitive normal in the end of last follow-up will be assessed by the proportional-odds cumulative (POC) logit model, commonly used for ordinal data. Under the proportional odds assumption, we assumed that the effect estimated between each pair of outcomes across two response levels are the same so we will test the proportional odds assumption for model appropriateness. Covariates such as SES, demographics, biomarker, imaging, and genetic risk factors for identifying observed transitions will be tested and evaluated in the model. Since there are a large number of variables will be involved in the model, we will apply Benjamini and Hochberg procedure to account for the multiple comparison issue to keep the type I error α=.05 (116). For assessing NIA-AA stage transitions over the study period (e.g., normal to SCI, normal to MCI, MCI to mild ADRD), we will use the ordinal logistic Markov chain transition model, an extension of the POC model, which considers intermediary transition for equal space data with more than two response variables with the assumption that the conditional distribution of the outcome at time point T only depends on the state at previous time point T-1. A stepwise regression with covariates sex, racial/ethnic group and age will be used to select the predictive covariates from longitudinal cognitive, fluid, genetic, and imaging biomarker results to predict the longitudinal decline and transition across NIA-AA stages. The final model will include all significant (p-value<.05) independent variables, interactions, and all pre-retained variables such as sex (to obtain the sex-adjusted odds ratios and 95% CI, along with risk differences) and age effect in cognitive impairment. We will vary covariance matrices to derive the best model with the lowest Bayesian Information Criterion (BIC). If models do not converge, we will use generalized estimating equation (GEE) with covariance matrices.

Understanding how risk factors work together and developing a predictive model for identifying high-risk individuals is one important aim of this proposal (117, 118). However, age is not only the strongest risk factor for ADRD but is also the strongest risk factor for death. Since the mortality rate in this cohort is not ignorable, we may consider a discrete time competing risk model via generalized additive regression. It is clear that the conversion risks from normal to MCI and from MCI to ADRD increases sharply after the age 80. Thus, once all the coefficients and functions of the regression models are estimated, we will construct the transition probability matrix considering age among the cognitively normal, cognitively impaired (MCI/ADRD), and at death. We will also test for back conversion from MCI/ADRD to normal and from ADRD to MCI. This probability matrix can then be used to estimate the future conversion risk for an individual as well as the future composition of normal, MCI, and ADRD participants in a group of participants. Cognitive progression models can be constructed by using the multi-state Markov models with two competing absorbing states: MCI/ADRD and death, this is the commonly used model with three states: normal, MCI/ADRD and death, so the transition probability matrix are permitted from normal to MCI/ADRD, MCI/ADRD to death, normal to death, and MCI/ADRD to normal (117, 118).

### Ethical considerations and declarations

HBI has been approved by the University of Miami Miller School of Medicine Institutional Review Board. Participants will provide informed consent at baseline and will be re-consented as needed with protocol changes.

### Timeline of the study

HBI began recruiting participants in 2022. As of September 21, 2023, 156 participants have completed their baseline visit and a growing number of participants are being seen for their first follow-up visit. Enrolled participants are expected to be seen annually until the point at which they display moderate to severe ADRD symptoms and are no longer able to complete the evaluations, at which point they will be discontinued from the study. HBI is designed to be an ongoing, longitudinal cohort that recruits at least 500 participants and then maintains that sample size by refreshing the cohort to counteract any participant drop out.

## DISCUSSION

HBI is planned as an ongoing, longitudinal observational cohort study of brain health among 500 racially/ethnically diverse participants with no or mild cognitive impairment living in the South Florida region. It is differentiated from other cohorts of brain health and ADRD in its deep phenotyping and collection of a host of novel predictors (e.g., neighborhood social determinants of health, epigenetics) of risk for and resilience against ADRD and cognitive decline measured over the life course, in characterizing and validating new ADRD (e.g., neurobehavioral) biomarkers, and in employing unique approaches and methods (e.g., machine learning, transdisciplinary, exposome, etc.). Standardized questionnaires and assessments used across multiple grant funded projects ensures comparability of measures and pooling of data that will permit high powered studies, which is particularly important for studies such as those examining effect measure modification that require large sample sizes. The cohort study will be significant in not only identifying modifiable factors that can be targeted via interventions to promote brain health and reduce ADRD risk, but also in its creation of a data and biospecimen repository, that can be shared in collaboration with the larger research community.

HBI’s limitations include a potential lack of generalizability of findings to certain communities and groups. Commonly, research volunteers tend to be more educated and less diverse than the general population and socioeconomically disadvantaged populations can be harder to retain in longitudinal cohort studies. Targeted recruitment efforts will be made to ensure a diverse group of participants are enrolled, including regular talks and events (e.g., health screenings) in communities that are representative of the diversity HBI aims to achieve. CCBH has begun building relationships with diverse communities in the South Florida area and plans to expand upon and support these relationships to develop an awareness of CCBH and to develop trust with local communities. Various retention efforts will be employed such as regular newsletters, telephone follow-up calls in between the annual visits, and sending birthday cards to participants. Transportation costs are covered for individuals who have no other means of traveling to CCBH to complete their HBI visits.

The expansive nature of the data collected at each annual visit places some burden on participants’ time, which could also affect retention. The CCBH team regularly weighs the utility of the data being collected in fulfilling HBI’s goals against participant burden. When needed, adjustments are and will be made to questionnaire forms, assessments, or the order of visit activities to minimize burden and/or incorporate new research questions.

The goal of recruiting and maintaining a steady cohort of 500 participants involves forethought. CCBH works to ensure it has the resources, staff, and scheduling time required to achieve this recruitment goal, including availability of ample facility spaces to conduct the visits and individuals who are cross-trained in multiple assessments to cover staff who are out of the office. The structure and timing of visits are designed to ensure the maximum number of participants can be seen over a given week, and a Clinical Trial Management System will be implemented to efficiently track all visits, reminder calls to participants, schedule staff time, supplies, and the like. New staff and additional research space will be acquired as needed to meet the recruitment sample size goals.

Findings from HBI will be disseminated broadly in a variety of formats. Results will be presented at academic conferences; at community events; and at local, national, and international seminars/presentations; and findings will be published in peer-reviewed journals. Newsworthy findings will be broadcast via press releases to the University of Miami media office and local and national print and other media sources. In addition, highlights from the study will be included in our regular newsletter for participants.

In summary, HBI is poised to make substantial contributions to the growing field of research on brain health and risk and resilience factors associated with ADRD. The data collected will lead to breakthroughs in developing new diagnostics and therapeutics, create comprehensive diagnostic evaluations, provide the evidence base for precision medicine approaches to dementia prevention with individualized treatment plans, provide educational and training opportunities, and create information clearinghouse activities for participants, families, caregivers, the lay public, health professionals, and policy makers.

## Author contributions

Dr. Besser: Conceptualization, Writing – Original Draft Preparation, Writing – Review & Editing; Dr. Chrisphonte: Writing – Original Draft Preparation, Writing – Review & Editing; Dr. Kleiman: Writing – Original Draft Preparation, Writing – Review & Editing; Dr. O’Shea: Writing – Original Draft Preparation, Writing – Review & Editing; Dr. Rosenfeld: Writing – Original Draft Preparation, Writing – Review & Editing; Dr. Tolea: Writing – Original Draft Preparation, Writing – Review & Editing; Dr. Galvin: Conceptualization, Writing – Original Draft Preparation, Writing – Review & Editing, Supervision.

## Data Availability

Data produced for the Healthy Brain Initiative will be available upon reasonable request to authors.

## Acknowledgements

The Healthy Brain Initiative is supported by NIH *R01AG071514, R01AG071514S1, R01NS101483, R01NS101483S1, RF1AG075901, and R56AG074889*. HBI would not be possible without the support of the Comprehensive Center for Brain Health’s staff and postdoctoral fellows: Katherine Almonte, Mirza Baig, Andrea Buehler, Yolene Caicedo, Simone Camacho, Gabrielle Czerwonko, Iris Cohen, Shanell Disla, Reem Ezzeddine, Conor Galvin, Gregory Gibbs, Keri Greenfield, Adolfo Henriquez, Mahesh Joshi, Elaine Le, Nicole Mendez, Claudia Moore, Aimee Ortiz, Mary Lou Riccio, Amie Rosenfeld, Ciny Singh, Monica Sonnenberg, Rebecca Smither, Marcia Walker, and Willman Zambrano. Dr. Besser is supported by NIH/NIA K01 AG063895, NIH/NIA R21 AG075291, and Alzheimer’s Association Research Grant (AARG-21-850963).

## Competing interests

All authors have completed the ICMJE uniform disclosure form at www.icmje.org/coi_disclosure.pdf and declare: that the submitted work is supported by NIH *R01AG071514, R01AG071514S1, R01NS101483, R01NS101483S1, RF1AG075901, and R56AG074889*; there are no financial relationships with any organizations that might have an interest in the submitted work in the previous three years; and no other relationships or activities that could appear to have influenced the submitted work.

## Notes

### Competing Interest Statement

The authors have declared no competing interest.

### Author Declarations

Ethics committee/IRB of University of Miami Miller School of Medicine gave ethical approval for this work.

